# Climate-driven spatiotemporal dynamics of *Aedes* infestation and dengue transmission in Porto Alegre, Southern Brazil

**DOI:** 10.64898/2026.03.31.26349860

**Authors:** Adryan Aparecido da Silva, Álvaro Gil Araujo Ferreira, José Lourenço, Amanda Cupertino de Freitas

**Affiliations:** Faculty of Pharmacy, Universidade Federal de Minas Gerais, Belo Horizonte, Minas Gerais, Brazil; Mosquitos Vetores, Instituto René Rachou – Fiocruz Minas, Belo Horizonte, Minas Gerais, Brazil; Universidade Católica Portuguesa, Católica Medical School, Católica Biomedical Research Centre, Oeiras, Portugal; Ecovec, Belo Horizonte, Minas Gerais, Brazil

## Abstract

Dengue transmission is strongly influenced by climatic conditions that affect mosquito population dynamics and virus circulation. In Southern Brazil, where dengue historically occurred at low levels, recent climatic anomalies may be contributing to the expansion of *Aedes* vectors and an increase in local dengue incidence. This study investigated the spatiotemporal association between climatic variables, *Aedes* mosquito infestation and dengue cases in Porto Alegre (Southern Brazil, 2018 to 2025). Entomological, surveillance and climatic data were analyzed using Moran’s I and LISA for spatial association, Kendall correlation, polynomial regression and LASSO to identify relevant drivers and develop predictive models of mosquito infestation and dengue incidence. A strong spatial association between *Aedes aegypti* and *Aedes albopictus* was observed, with persistent local clusters detected across all years. Annual climatic variables were associated with mosquito abundance in several districts. Overall, rainfall frequency had a stronger effect on *Aedes aegypti* abundance than accumulated rainfall. Temperature and lagged infestation indices showed strong association with both species and dengue incidence, with effects observed up to four weeks prior. Predictive models demonstrated good agreement between observed and predicted values, particularly within low to moderate infestation levels. Lagged variables were consistently retained in both mosquito infestation abundance and dengue incidence models, highlighting the importance of temporal predictors for anticipating vector dynamics and dengue risk. This approach is generally applicable for predicting *Aedes* infestation and disease incidence and emphasizes the importance of integrating entomological and climatic surveillance data to improve anticipation and detection of dengue risk periods and support more effective public health interventions.

**Author summary:** Dengue is a viral disease transmitted by mosquitos that affects millions of people worldwide. Its spread is strongly influenced by climatic conditions that modulate mosquito life cycles. In Southern Brazil, where dengue historically occurred at low levels, recent climate variability and extreme weather events may be creating favorable conditions for the expansion of *Aedes mosquitos* and local dengue activity. In this study, we analyzed mosquito, dengue, and climatic surveil in Porto Alegre, Brazil, from 2018 to 2025 to understand how climate influences mosquito infestation and dengue incidence. We found that mosquito infestation and dengue cases show clear spatial patterns across the city and are strongly influenced by temperature and rainfall. Interestingly, the number of rainy days had a stronger effect on mosquito abundance than the total amount of rainfall. We also observed that higher mosquito infestation was associated with an increase in dengue cases up to four weeks later. These findings show that combining climatic and mosquito surveillance data can help anticipate periods of higher dengue risk and support more effective mosquito control and public health actions.

## Introduction

Dengue, one of the most important mosquito-borne diseases globally, is well documented in tropical and subtropical areas, causing outbreaks in South America, Southeast Asia and the Western Pacific [1,2]. However, recent events associated with climate change, globalization and urbanization have shifted its geographic expansion towards broader latitudes [3,4].

Mosquito reproduction, longevity and competence for viral transmission depend on climatic conditions [5–8]. *Ae. aegypt*i and *Ae. albopictus,* the key vectors of dengue, have a short life cycle, with full development from egg to adult within a couple of weeks [9]. Consequently, their population can be affected both by long-term seasonal weather fluctuations (e.g. temperature, humidity) or short-term extreme events such as floods, heavy rains or heat waves [10,11].

*Ae. aegypti* was historically restricted to the African continent, whereas *Ae. albopictus* to Southeast Asia. Both species have undergone major geographic expansion over the past century, largely driven by globalization processes (e.g. increased human mobility) and environmental changes that create new suitable habitats [12,13]. Currently, *Ae. aegypti* shows a near-global distribution and is considered the primary dengue vector in urbanized regions [14–16]. *Ae. albopictus*, in turn, is recognized for its greater ecological plasticity and tolerance to a wider range of climatic conditions, been reported in Brazil for the first time in 1986 in Rio de Janeiro and Minas Gerais (Southeast) [17,18].

In 2024, Brazil reported near 6.5 million suspected dengue cases, representing almost half of the cases reported in the Americas [19,20]. The virus is well-distributed across the entire country with all four serotypes identified indicating hyperendemicity [21,22]. The current dengue epidemiological scenario in Brazil presents a unique challenge, as predicting local transmission relies on the complex interplay between human, climatic and mosquito factors [23–25].

In the first 3 months of 2023, 63 municipalities detected local dengue transmission for the first time, half of them in Southern Brazil. This region has historically been associated with low *Aedes* infestation and dengue incidence, but in recent years has witnessed both mosquito and dengue activity. Recent evidence suggests that environmental change may create suitable conditions for both mosquitos and viruses, which may be facilitating emergence in Southern Brazil [22–24].

Despite growing evidence linking climatic conditions to mosquito population dynamics and dengue transmission, important knowledge gaps remain, particularly in temperate regions where dengue has historically been limited. Understanding how climatic variability influences vector infestation and disease dynamics at the local scale is essential for improving surveillance and early warning systems.

Porto Alegre, the capital city of Rio Grande do Sul, the most Southern state in Brazil, has faced extreme weather events in recent years, including temperature anomalies in 2023, flooding in 2024 and heavy rains in 2025. This study investigates the relationship between climatic variability, mosquito infestation and dengue incidence in Porto Alegre between 2018 and 2025 through an integrated analysis of entomological, epidemiological and climatic surveillance data.

## Methods

### Study region

Porto Alegre is one of the main Brazilian’s metropolises, located in the Southern Brazil, with 1,388,794 inhabitants (2022). It covers an area of 495.39 km² with nearly 50% of urbanization. Climatic conditions are temperate climate with well-defined seasons.

### Entomological data

Data were obtained from MosquiTRAPs as part of the municipal surveillance system in Porto Alegre. Trap installation was made in strategic locations based on local dengue cases and mosquito infestation (S1 Fig). Numbers of female *Aedes* adults were assessed weekly and RT-PCR was performed to detect dengue virus (DENV) serotypes (DENV1-4) in a bulk of adult mosquitos collected. All infestation data were normalized by the number of traps installed in the same period. The DENV-positive trap index was calculated as the number of traps with dengue virus detection divided by the total number of traps installed per year × 100. The Mean Female *Aedes* Index (MFAI) was calculated as the number of adult female *Aedes* mosquitos captured per week divided by the total number of traps inspected in the same week.

### Climatic data

The climatic data were sourced from the Copernicus Data Store [26], a repository maintained by the European Centre for Medium Range Weather Forecasts (ECMWF) with daily resolution for the time zone Etc/GMT-3 and based on Porto Alegre’s spatial location. Air temperature at 2 m above ground level and dew point were converted to Celsius by subtracting 273.15. For each week and year, aggregated values were obtained by the daily observations and subsequently calculating the mean. Relative humidity was obtained using the formula:

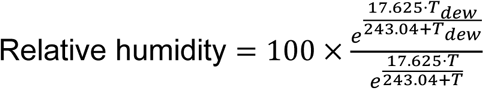

Precipitation was converted to millimeters and aggregated to obtain accumulated per week and year. For accumulated days with precipitation analysis, 1mm of rainfall was established as the threshold for rainy days.

### Viral-mosquito climatic suitability

To evaluate climatic conditions that promote transmission, the climate-driven mosquito-viral suitability Index P was obtained from a free-to-use global synthetic dataset [27]. To assess the joint effect of mosquito-viral suitability and vector infestation, we created the Vector-Fitness Infestation Index as:

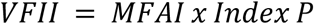

### Land cover data

For the assessment of land cover in the study area, data were obtained from Mapbiomas Brasil [28]. Each raster was downloaded and reprojected to match the coordinate reference system of the Porto Alegre municipal boundary shapefile. Subsequently, the rasters were cropped and masked to the Porto Alegre study area to retain only pixels within the municipality limits. The resulting rasters were converted to tables containing pixel coordinates and land cover class codes for each year. Land cover classes were then grouped into two broader categories: anthropic land uses (urbanized areas, pasture, agriculture, rice, sugarcane, mining, and other land uses) and vegetation cover (forest formation, savanna formation, grassland formation, and planted forest), according to the MapBiomas classification scheme.

Because anthropic land use and vegetation cover changed minimally during the study period, these variables were not included in the statistical analyses (S2a–b Fig).

### Dengue cases

Dengue is a notifiable disease in Brazil and cases are reported to the Notifiable Diseases Information System (SINAN) of the Brazilian Ministry of Health. Cases are initially notified based on clinical evaluation of symptoms, while a subset is confirmed by laboratory diagnosis.

Incidence data from suspected dengue cases between 2018 and 2025 were obtained from InfoDengue [29], an open Brazilian surveillance platform that integrates cases from SINAN. The information of all autochthonous cases was retrieved from the official Power BI epidemiological dashboard maintained by the Municipality of Porto Alegre [30].

### Statistical analysis

#### Spatial autocorrelation

For each year, a spatial analysis of *Ae. aegypti*, *Ae. albopictus*, DENV+ traps index and dengue cases was performed using Moran’s Global Index (Moran’s I) [31] and Local Indicators of Spatial Association (LISA) to assess the geographical association and clustering in the study area. To perform the tests, a neighborhood matrix was created using k-nearest neighbors (KNN). For each unit, the four nearest neighbors were identified based on Euclidean distances between centroid coordinates.

Positive values of Moran’s I indicate spatial clustering, whereas values close to zero suggest a random spatial pattern and negative ones indicate spatial dispersion in the area. Statistical significance was determined using a permutation test.

LISA identifies local clusters by classifying each spatial unit into High-High or Low-Low, indicating areas surrounded by similar values (clusters), and High-Low or Low-High indicating areas above average surrounded by areas below average (spatial outliers). While Moran’s I provides a global measure of spatial correlation, LISA focuses on the identification of specific locations driving spatial dependence.

### Correlation and Modeling

#### Kendall correlation

Associations between climatic variables, entomological indicators and dengue incidence were evaluated using Kendall’s rank correlation coefficient (τ). To explore local variability in climate-entomology relationships across the area per year, Kendall correlation was calculated separately for each district. For increased temporal resolution, a weekly correlation was applied. Considering potential delayed effects, lagged variables ranging from zero to four weeks were also included.

#### Polynomial regression

To capture the effect of rainfall frequency on mosquito infestation, 1 mm was established as a rainy occurrence and 5, 10 and 30 days as distinct time-windows representing short-, medium- and long-term rainfall persistence. For each epidemiological week, the MFAI was compared with the rainfall frequency derived from the preceding period. Polynomial regression models were then fitted from first to fourth order to evaluate the non-linear relationship between variables. The optimal model was selected based on the Akaike Information Criterion (AIC), with lower values indicating better model fit while accounting for model complexity. Additionally, analysis of variance (ANOVA) was performed to assess if increases in polynomial degree resulted in a statistical improvement of performance. Second order polynomial was best for a 5 and 10-day period, while a fourth-order polynomial was best for a 30-day period.

#### LASSO

To model the association between climatic variables, entomological indicators and dengue incidence, Least Absolute Shrinkage and Selection Operator (LASSO) regression was employed. This method introduces a penalty that shrinks less informative variables to zero, reduces model complexity and mitigates multicollinearity. Three LASSO models were fitted: two models to address weekly mosquito infestation based on lag climatic variables and a third to evaluate the joint effect of Index P, MFAI and VFII of both species on weekly log (dengue incidence + 1).

The lambda value that minimized the cross-validated error was used to penalize the coefficients. A Gaussian distribution was assumed and a ten-fold cross-validation was applied to train the model. Its performance was evaluated using the root mean squared error (RMSE), compared to a Linear Model, and the deviance ratio over the intercept-only model.

All analyses were conducted using R (version 4.4.1). Spatial analysis was performed using the R package *spdep.* LASSO analysis, Kendall and polynomial regression were performed with the *glmnet* and *stats* R packages, respectively.

## Results

### Porto Alegre spatio-temporal *Aedes* surveillance from 2018 to 2025

Across the period, around 50 districts remained under surveillance (S3a Fig). 1,447 distinct traps were installed, capturing 130,122 female *Ae. aegypti* and 4,618 *Ae. albopictus* over 375,776 inspections. *Aedes* represented the dominant genus among all mosquitos collected (S4 Fig). Weekly inspections increased in 2019, reached an average of 1,400 inspections per week, decreasing during 2020 and remaining stable until 2025 (Fig 1a). MFAI data indicated an increase in infestation since 2021 (Fig 1b). Both absolute counting and MFAI values were significantly higher for *Ae. aegypti*, demonstrating lower capture levels of *Ae. albopictus in* Porto Alegre.

**Fig 1.**
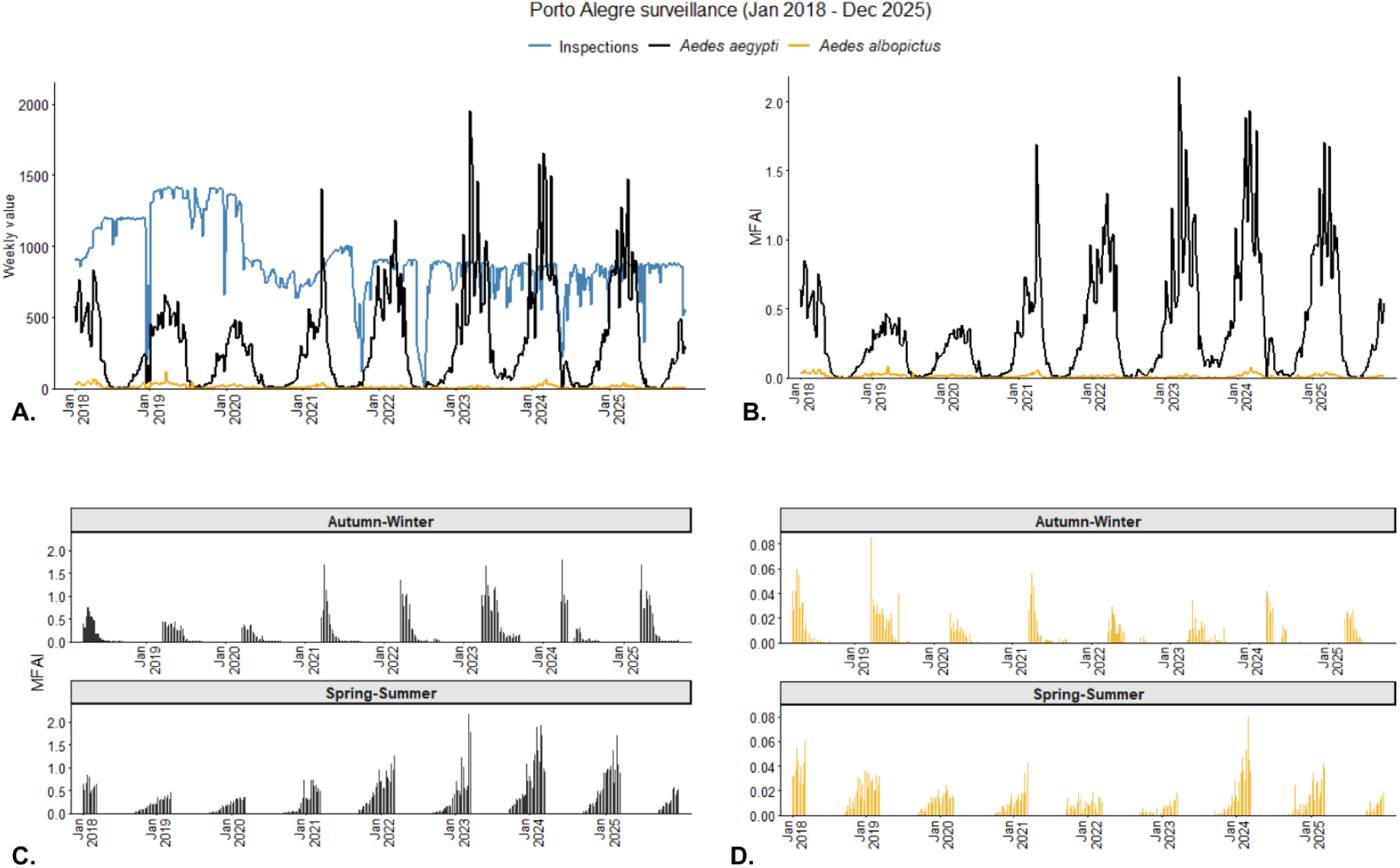
Temporal dynamics of mosquito surveillance indicators in Porto Alegre, Brazil, from 2018 to 2025. **(a)** Weekly values for inspections, *Aedes aegypti* and *Aedes albopictus* in Porto Alegre. **(b)** Weekly values of MFAI for both species. **(c)** MFAI of *Aedes aegypti* in Autumn-Winter (upper) and Spring-Summer (lower). **(d)** MFAI of *Aedes albopictus* in Autumn-Winter (upper) and Spring-Summer (lower).

For *Ae. aegypti,* the highest number of captured females was observed during spring-summer (Fig 1c): peaking 1,951 in the epidemiological week 11 in 2023 (March). While weeks 31 in 2021 and 2022 (July), 32 and 33 in 2022 (August) and 21 in 2024 (April), all in autumn-winter seasons, were the only ones with no captures. On the other hand, three of the four weeks with the highest capture of *Ae. albopictus* occurred in autumn-winter (Fig 1d): epidemiological week 13 in 2019 (March) with 119 females captured, followed by week 16 and 17 in 2018 (April) with 62 mosquitos trapped. The highest level in the spring-summer period was observed in week 10 in 2024 (March), with 69 females captured.

The number of DENV-positive traps increased steadily after 2021, reaching 122 positive traps in 2025 (S3a Fig). When accounting for sampling effort, the annual positivity rate, calculated as the proportion of DENV-positive traps among traps with mosquito capture, remained low overall but increased over time, rising from 0.01% in 2018 and 0.02% in 2019 to 0.46% in 2022, 0.33% in 2023, 0.40% in 2024, and peaking at 0.98% in 2025, indicating that, despite low overall detection rates, viral circulation intensified over time. Pools of trapped mosquitoes were tested for serotype identification, indicating cocirculation of DENV-1 and DENV-2 in 2023 and 2024 (S3b Fig).

The spatial analysis of infestation was conducted to detect spatial autocorrelation and clusters of *Ae. aegypti* and *Ae. albopictus* using the ratio of the total female captures by the total traps installed per year. Spatial autocorrelation peaked in 2023 for both species (S1 Table), with a marked strong correlation for *Ae. aegypti* yearly.

In 2018 *Ae. aegypti* was concentrated in Partenon, Northwest, Central-South, Northeast and Cruzeiro regions, spreading over different areas until 2025. A notable clustering in the North, East and Eixo Baltazar regions in recent years was observed, with its proportion reaching more than 40 mosquitos per trap (Fig 2a). Despite the presence of clusters in other areas across all years, *Ae. albopictus* seems to be restricted to the South and Central-South regions, reaching up to 6 mosquitos per trap in 2019 and 2024 (Fig 2b). This was consistent with the higher capture rates observed for *Ae. aegypti* in Porto Alegre.

**Fig 2.**
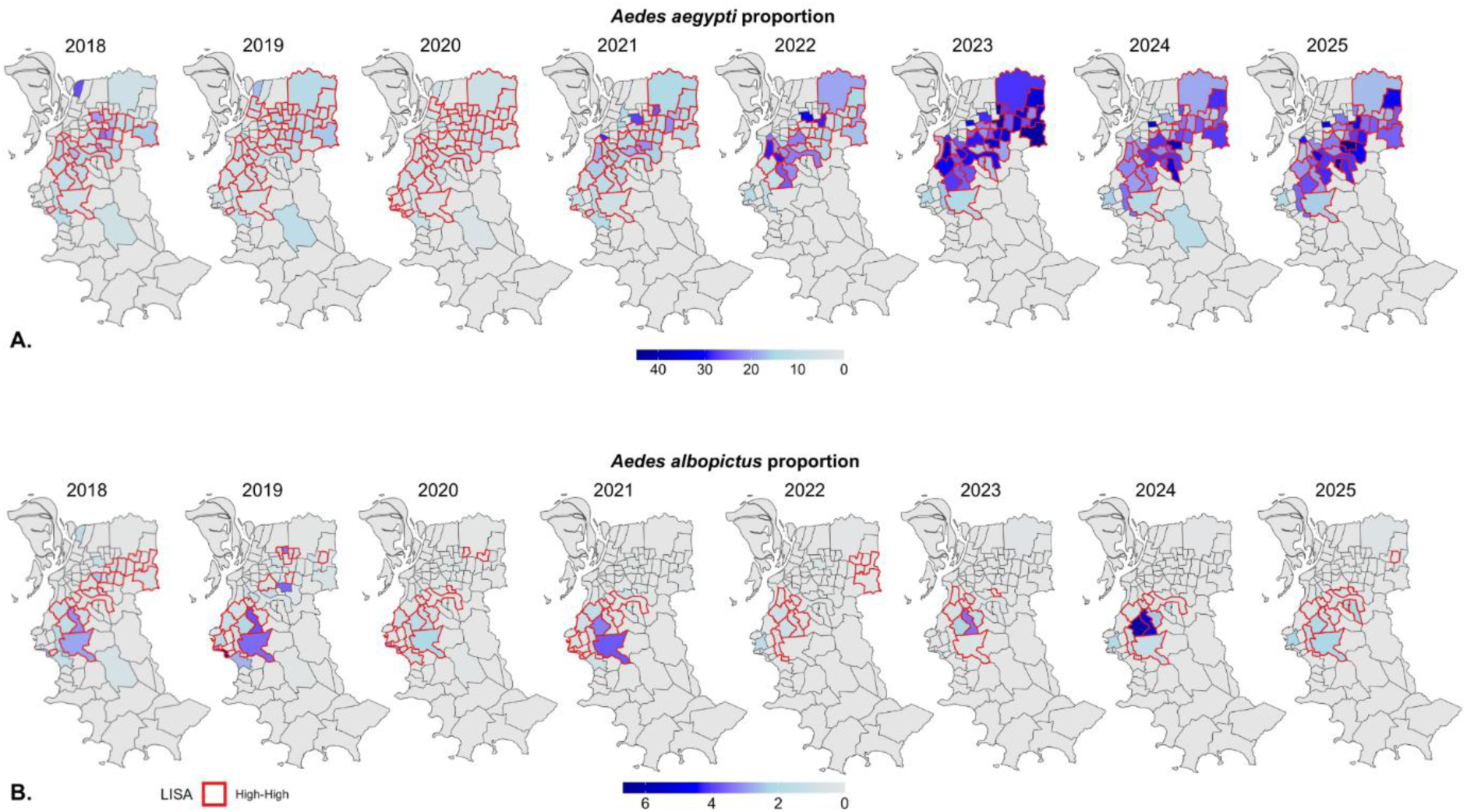
Spatiotemporal clustering of *Aedes aegypti* and *Aedes albopictus* proportions in Porto Alegre, Brazil, from 2018 to 2025. **(a)** Spatial clustering of *Aedes aegypti* in Porto Alegre from 2018 to 2025; *Ae. aegypti* proportion represents the total mosquitos captured by the total traps each year**. (b)** Spatial clustering of *Aedes albopictus* in Porto Alegre from 2018 to 2025; *Ae. albopictus* proportion represents the total mosquitos captured by the total traps of each year. Red lines around districts indicate high-high LISA clusters, representing high values surrounded by areas with similarly high values.

### Climatic factors influence mosquito dynamics in Porto Alegre

Porto Alegre has a temperate climate, with well-defined seasons, weather and climate conditions. Each climate variable exhibited different effects across districts. Accumulated precipitation had a spatial variance between April and June, with rainfall ranging from 10 to 100 mm, and between September and December, reaching 200 mm in some regions (Fig 3a).

**Fig 3.**
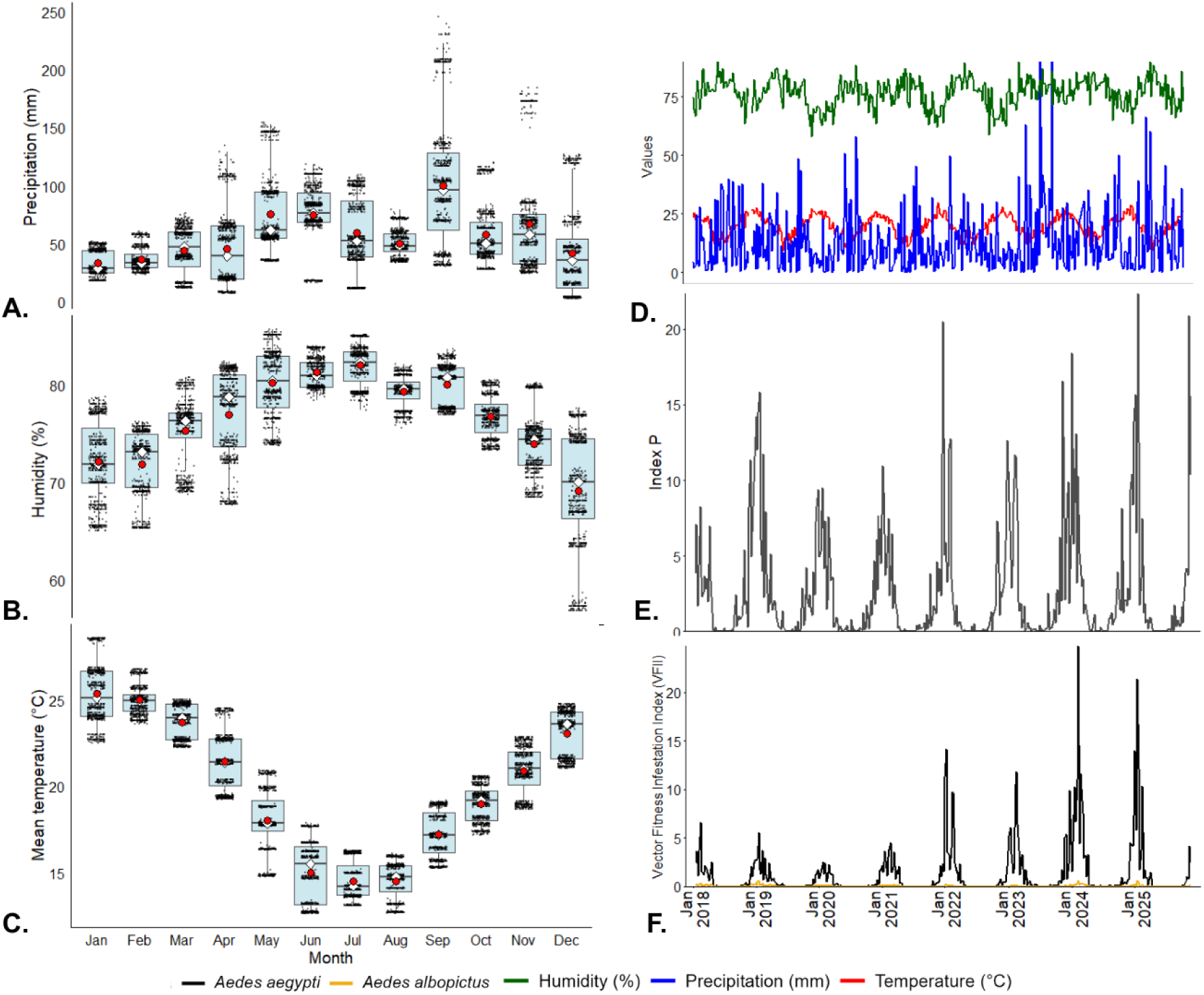
Climatic variability and vector suitability indices associated with mosquito dynamics in Porto Alegre, Brazil, from 2018 to 2025. Monthly distribution of precipitation (mm), relative humidity (%), and mean temperature (°C) across districts. Each point represents the mean value per district across all years; red circles indicate the mean and white diamonds the median**. (D)** Weekly time series of relative humidity, mean temperature, and precipitation. **(E)** Weekly Index P derived from temperature and humidity. **(F)** Weekly Vector Fitness Infestation Index (VFII) for both species.

For relative humidity, December was the month with the highest variance, with a range from 50% to 75%, followed by January to May (Fig 3b). Mean temperature showed lower spatial variability, although January, April, May, and June showed the highest temperature variance within the study area (Fig 3c). Critically, the winter months (June to August) showed temperature ranges below 15°C, which is considered a critical biological threshold for mosquito life cycles [8].

In the epidemiological weeks 36 and 46 (September and November) in 2023, the rainfall reached approximately 100 mm. The highest mean temperature values occurred in 2022 and 2025, with nearly 30°C recorded in January and February, respectively. The highest humidity levels were observed between May and July of each year, reaching nearly 90% (Fig 3d).

Index P values indicated climatic suitability for transmission. In the study area, suitable conditions for female *Aedes* were estimated, broadly, between October and May (late spring to early autumn). Since 2023, the Index P has increased seasonally (Fig 3e).

To further explore the effect of mosquito infestation and climate-driven transmission potential, the MFAI and Index P values were combined to create the Vector Fitness Infestation Index (VFII). VFII showed the greatest potential from September to April each year. The highest values were reported in epidemiological week 6 of 2025 (February), where VFII reached 24 and 0.59 for *Ae. aegypti* and *Ae. albopictus* (respectively), followed by week 3 in 2025 (January) with 21 and 0.61 for *Ae. aegypti* and *Ae. albopictus*, respectively (Fig 3f). Although the Index P did not discriminate between species, the lower infestation of *Ae. albopictus* resulted in much lower VFII compared to *Ae. aegypti*. The already reported increases in index P over time became more apparent from VFII. A linear regression analysis revealed a significant positive temporal, by week trend in the VFII for *Ae. aegypti* (β = 0.0038, p = 0.001), indicating a gradual increase in transmission potential due to climatic suitability over the study period. In contrast, no significant temporal trend was observed for *Ae. albopictus* (β = −0.00007, p = 0.058).

Kendall correlation analysis revealed multiple associations between background variables and mosquito species. A strong association between climatic variables and *Aede*s infestation over the period in different districts (S2 Table). Boa Vista, Jardim Itu, Jardim Leopoldina, Jardim Sabará, Vila Jardim, Três Figueiras and Mário Quintana had a strong association of accumulated precipitation with *Ae. aegypti.* Três Figueiras also showed a correlation with relative humidity. Partenon and Sarandi had an association of relative humidity and accumulated precipitation with *Ae. albopictus*, respectively. In Teresópolis and Glória, both increase in relative humidity and mean temperature were associated with an increase in *Ae. albopictus* infestation.

To verify the effect of lagged climatic variables on mosquito dynamics, weekly values of accumulated precipitation, mean temperature and relative humidity were correlated with MFAI of both species. The temperature was the only one positively correlated, with a gradual increase over time. Humidity had a negative effect on the infestation index, and weekly accumulated precipitation seems to barely influence the mosquito presence (Table 1).

**Table 1.** Kendall correlation of weekly lagged climate variables and MFAI. *** p<0.001.

| Weeks<br>lag | $\tau$ | | | | | |
| --- | --- | --- | --- | --- | --- | --- |
|  | MFAI - *Aedes aegypti* |  |  | MFAI - *Aedes albopictus* |  |  |
|  | Precipitation | Temperature | Humidity | Precipitation | Temperature | Humidity |
| 0 | -0.093*** | 0.419*** | -0.2158** | -0.1066*** | 0.4048*** | -0.1991*** |
| 1 | -0.1245*** | 0.4789*** | -0.2339** | -0.1106*** | 0.4661*** | -0.1986*** |
| 2 | -0.1033*** | 0.5049*** | -0.2518** | -0.1259*** | 0.4857*** | -0.2180*** |
| 3 | -0.0993*** | 0.5483*** | -0.2681** | -0.0734*** | 0.5056*** | -0.2323*** |
| 4 | -0.094*** | 0.5809*** | -0.2930** | -0.0852*** | 0.5352*** | -0.2547*** |

The relationship between the number of rainy days and mosquito MFAI indicated a main effect on *Ae. aegypti,* peaking at approximately four days out of 5 (Fig 4a) and at approximately eight days out of 10 (Fig 4b). In a 30-day window, the infestation suggests a non-linear pattern, with limited predictive accuracy, increasing at 23-24 rainy days followed by a slight decline (Fig 4c). *Ae. albopictus* did not exhibit an association with the number of rainy days in any of the evaluated time windows. The low infestation levels of this species in the study area may have reduced the ability to detect consistent environmental relationships.

**Fig 4.**
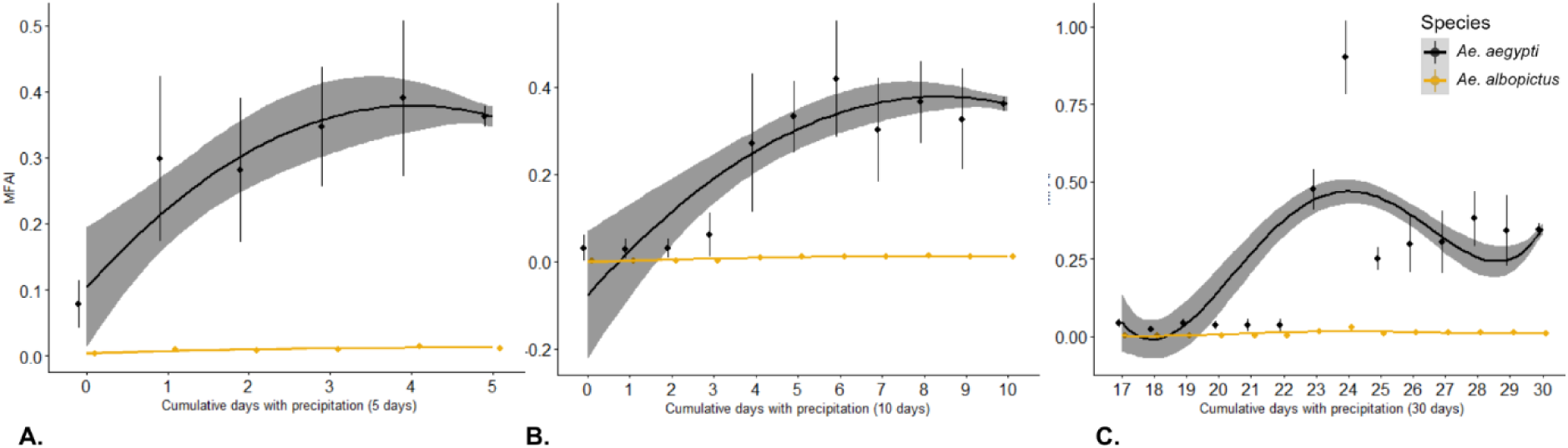
Nonlinear relationship between cumulative rainy days and mosquito abundance (MFAI) for *Aedes aegypti* and *Aedes albopictus*. Association between MFAI and the cumulative number of rainy days over 5-day **(A)**, 10-day **(B)**, and 30-day **(C)** periods. Points represent mean values and error bars indicate standard errors. Solid lines represent polynomial regression fits, with shaded areas indicating 95% confidence intervals. Second-order polynomials provided the best fit for the 5- and 10-day periods, whereas a fourth-order polynomial best described the 30-day period.

### Mosquito infestation modulates dengue dynamics in Porto Alegre from 2018 to 2025

From 2022 to 2025, the dengue incidence per 100,000 inhabitants has constantly increased (S5 Fig). Notably, the epidemiological weeks 13 to 21 in 2025 (March to May) reached the highest incidence levels across the period.

The first detection of autochthonous dengue cases was reported in 2010 in Jardim Carvalho (East Region). Since then, the disease has spread across the city, with diagnoses made based on clinical symptoms, and a percentage of these confirmed through laboratory analysis. Between 2019 and 2025, 72,965 dengue cases were reported, 51% of which were confirmed and 93% of these were classified as autochthonous. In 2025, Passo das Pedras district (Eixo Baltazar region) reported the highest burden of autochthonous cases, with 2,364. Followed by Rubem Berta district (Eixo Baltazar region) with 2,236 cases, a weak spatial autocorrelation was detected (S3 Table). In 2024, Restinga (Restinga region) and Partenon (Partenon region) reported 1,171 and 1,029 autochthonous cases, respectively. Partenon also reported a high number of cases, with 1,377 registered across 2023, while in 2022, Jardim Carvalho registered 1,039 cases (S6a-d Fig).

Between 2022 and 2025, some districts exhibited localized hotspots where up to 50% of inspected traps tested positive for dengue virus (S6e–h Fig). However, overall positivity remained low when considering the total number of inspections, indicating that viral circulation was spatially heterogeneous and concentrated in specific areas rather than widespread across the city. Spatial autocorrelation of DENV-positive traps increased over time, with moderate clustering observed in 2023 and peaking in 2025 (S4 Table), coinciding with districts reporting the highest number of autochthonous dengue cases. Additionally, Kendall correlation analysis supported a positive association between *Ae. aegypti* infestation (MFAI) and autochthonous dengue cases (S5 Table).

To assess the predictive potential of MFAI data for dengue incidence, Kendall correlation analysis using lagged weekly MFAI values was applied to assess its relationship with dengue incidence. The coefficients increased up to 4 weeks, indicating a progressively stronger temporal association and suggesting a lead relationship of mosquito infestation on dengue occurrence (Table 2).

**Table 2.** Kendall correlation - Lagged MFAI and autochthonous dengue cases. *** p<0.001.

| Weeks lag | $\tau$ | |
| --- | --- | --- |
|  | MFAI - *Aedes aegypti* | MFAI - *Aedes albopictus* |
| 0 | 0.2737*** | 0.1206*** |
| 1 | 0.3376*** | 0.322*** |
| 2 | 0.3911*** | 0.3645*** |
| 3 | 0.4470*** | 0.4085*** |
| 4 | 0.4953*** | 0.4479*** |

### Modeling entomological infestation and dengue incidence

Predictive models were applied to evaluate the contribution of climatic and entomological variables to mosquito infestation and dengue incidence. For *Ae. aegypti* infestation, a linear model (LM) produced a cross-validated RMSE of 0.2866, whereas a LASSO model achieved 0.2864. The LASSO deviance ratio was 0.52. For *Ae. albopictus*, RMSE values were 0.00977 for the LM and 0.00975 for the LASSO model, with a deviance ratio of 0.49. Although improvements in predictive error were minimal, penalization reduced the magnitude of coefficients among correlated climatic variables and improved model parsimony. Temperature at a three and four-week lags received the largest standardized penalized coefficient for *Ae. aegypti* and *Ae. albopictus*, respectively, indicating the strong contribution of temperature to model prediction among retained climatic variables (S6 Table).

For dengue incidence, the LM model produced an RMSE of 1.003, whereas the LASSO model produced an RMSE of 1.006. The deviance ratio was 0.61, indicating that 61% of null deviance was explained by the penalized model. MFAI of both species and VFII of *Ae. albopictus* obtained the highest coefficients, reinforcing the strong predictive relevance of mosquito infestation index (MFAI) (S6 Table).

Compared to the unpenalized linear model, LASSO produced smaller coefficient magnitudes, indicating shrinkage of potentially inflated estimates due to multicollinearity and improving model stability and generalizability. The model demonstrated a good predictive performance for *Ae. aegypti* MFAI, whereas predictive accuracy for *Ae. albopictus* was limited to lower infestation levels (Fig 5a, b). For the dengue model, prediction accuracy closely matched the log-transformed incidence values across the observed range (Fig 5c).

**Fig 5.**
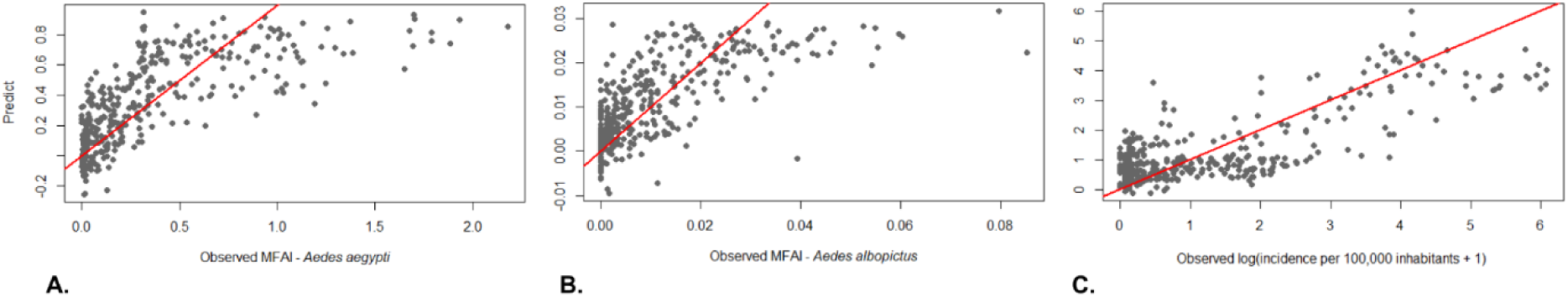
Predictive performance of LASSO regression models for mosquito abundance and dengue incidence. LASSO plots for observed versus predicted values of **(a)** MFAI – *Aedes aegypti* (RMSE = 0.2864, deviance ratio = 0.5225), **(b)** MFAI – *Aedes albopictus* (RMSE = 0.0097, deviance ratio = 0.4933) and **(c)** log(dengue incidence +1) (RMSE = 1.005, deviance ratio = 0.61. Red lines represent identity (y = x).

## Discussion

This study examines how climatic variables drive *Aedes* infestation and influence dengue cases in Porto Alegre, Brazil. Results confirm persistent spatial clustering of *Ae. aegypti* from 2018 to 2021, as previously reported [32] expansion into new districts between 2022 and 2025 in the Northwest, North, Cruzeiro and Central-South regions. In 2023, the year with the strongest spatial autocorrelation (S1 Table), several areas registered approximately 40 mosquitos per trap, corresponding to the highest weekly infestation index observed in the study. Notably, this coincided with the most pronounced expansion of dengue towards Southern Brazil.

Gloria, South, Central-South, Cristal and Cruzeiro emerged as critical areas for *Ae. albopictus* infestation, with persistent clusters across all years. In the Central-South, districts such as Nonoai and Teresópolis, characterized by lower population density, likely provide microhabitats and peridomestic breeding sites that favor the ecological profile of this species [33,34]. In 2025, an increase in infestation was also observed in the Partenon region, particularly in Vila São José and Vila João Pessoa, areas marked by high population density, informal settlements, and substantial peridomestic garbage accumulation [35]. These socioenvironmental conditions may create peri-urban-like niches within densely populated settings, highlighting the ecological plasticity of *Ae. albopictus* and its characterization as a predominantly peri-urban vector capable of adapting to urban environments where suitable breeding opportunities are present.

Annual analyses reinforced the influence of climatic variables at the district level, with 3 of the most populated Porto Alegre’s districts (Mario Quintana, Partenon and Sarandi), showing strong correlation of accumulated precipitation and relative humidity to both *Aedes* species, while temperature was correlated with *Ae. albopictus* in Teresopolis and Gloria.

Regarding the influence of weekly climatic variables on mosquitos, temperature showed an increasingly strong association with *Aedes* abundance over time. The mosquito traits relevant to dengue transmission peak between 23 and 34°C [36] and temperature below 18°C are expected during winter in most of Southern Brazil. However, high MFAI was observed also in Autumn, revealing the maintenance of the vector reproduction cycle and suggesting its adaptation to lower temperatures, allowing the sustainability of disease transmission throughout the entire year [37,38].

Weekly relative humidity exerted a weak negative correlation, while accumulated precipitation barely affected population growth. In contrast, the relationship between the number of rainy days and *Ae. aegypti* infestation revealed a positive association within 5 and 10 days. Similar results have been reported in other temperate city [39], where consecutive rainy days likely maintain breeding sites stability and boost aquatic development, while disruptive rainfall may flush larvae and eggs from containers, reduce adult flight activity, and impair the dispersion of oviposition attractants, thereby decreasing trap captures [40,41]. Taken together, these results indicate that annual climatic variables reflect broader environmental suitability, whereas weekly weather fluctuation may alter short-term infestation dynamics.

Dengue local transmission relies on a complex interplay between climatic variables, herd-immunity, human movements and exposure, vector and DENV presence. An increase in mosquito density does not immediately translate into dengue cases, as the virus must complete its extrinsic and intrinsic incubation period before symptom onset. Additionally, delays in healthcare seeking system and case reporting likely explain the progressive correlation observed between mosquito infestation and autochthonous dengue cases [42]. *Ae. aegypti* and *Ae. albopictus* are also responsible for transmission of Zika virus (ZIKV) and Chikungunya virus (CHIKV) in Porto Alegre [43]. Nevertheless, CHIKV and ZIKV incidence were minimal, with sporadic cases reported (S7a,b Fig). This can either be explained by insufficient introduction of those viruses to spark outbreaks, or perhaps the lower infestation of *Ae. Albopictus* determines their lower potential for transmission [43].

Since 2022, VFII has increased seasonally for *Ae. aegypti,* indicating mosquito-viral capacity for disease transmission in these periods. There were observed peaks in 2024, when Brazil registered more than 6 million dengue cases [44], the highest number on record, and in 2025, the year with the highest incidence level in Porto Alegre. Further analysis should include the joint effect of mosquito infestation and viral transmission suitability on reported dengue incidence in Porto Alegre and elsewhere.

The application of LASSO predictive model allowed to integrate multiple variables and assess its joint effect on mosquito infestation and dengue incidence, reducing collinearity and shrinking variables to zero. Previous work reported its performance considering only precipitation variables on *Aedes* abundance [45]. In this study, LASSO was extended to consider weekly lagged mean temperature, accumulated precipitation, and relative humidity effects on MFAI. A pattern was observed in the prediction of residuals in all models: low observed values were frequently overestimated, whereas high values were consistently underestimated. This compression toward the mean likely reflects the combined effects of model linearity and coefficient shrinkage under penalization. LASSO regression minimizes overall prediction error relative to the intercept-only model, which encourages stabilization around central tendencies rather than precise fitting of extreme observations. Additionally, extreme infestation and incidence levels (i.e. peaks and troughs) may depend on variables not included in the present modeling framework. Consequently, while the selected lagged climatic and entomological predictors capture the dominant temporal structure of transmission dynamics, additional factors may be required to accurately reproduce extreme-magnitude events.

Interestingly, the MFAI of *Ae. albopictus* presented a strong negative penalized coefficient (S6 Table), whereas its VFII showed a positive one in the dengue incidence model, although with a smaller magnitude. This may reinforce the previous result that, *Ae. albopictus* infestation alone may be more prevalent in peri-urban contexts with lower dengue transmission, while its ecological fitness under favorable climatic conditions enhances its epidemiological relevance. These findings indicate the importance of distinguishing between mosquito presence and climate-driven viral transmission potential when modeling transmission dynamics and suggest that climatic suitability enhances the epidemiological relevance of *Ae. albopictus* in the study area.

Predictive models combining weekly climate and entomological indicators may serve as an early-warning component for disease surveillance systems and provide a powerful framework for public health planning for outbreak mitigation. Such integration can support targeted mosquito control interventions, optimize resource allocation across districts, and enhance preparedness during years of heightened climatic suitability such as those associated with unusually warm or persistent rainfall.

Taken together, these findings highlight the importance of integrating climatic information with routine vector surveillance to better understand and anticipate dengue transmission dynamics in temperate urban settings. As dengue continues to expand into regions historically considered low risk, such as southern Brazil, strengthening integrated surveillance systems will be essential for improving early detection of transmission risk and supporting timely public health responses in the context of ongoing climatic and environmental change.

## Data Availability

All relevant data are included within the manuscript and its Supporting Information files. Publicly available data were obtained from the InfoDengue platform (https://info.dengue.mat.br/) and the Copernicus Climate Data Store (https://cds.climate.copernicus.eu/). Entomological data from MosquiTRAP surveillance were obtained under institutional agreements with the Municipal Health Department of Porto Alegre and are not publicly available. Data can be made available from the corresponding author upon reasonable request and with permission from the relevant authorities.

## Acknowledgments

The authors thank the Municipality of Porto Alegre, Ecovec, and Rentokil for providing access to data and supporting this study. We also acknowledge the Parque Tecnológico de Belo Horizonte (BHTEC) for its support and collaboration. This publication was supported by the Fundação de Amparo à Pesquisa do Estado de Minas Gerais (Fapemig) – Grant APQ-04991-23 and the Instituto René Rachou/Fiocruz Minas. J.L. acknowledges the Fundação para a Ciência e Tecnologia (FCT) for institutional support to his host institution, CBR (DOI:10.54499/UID/06497/2025, UID/06497/2025).

## Supporting information

**S1 Fig.**
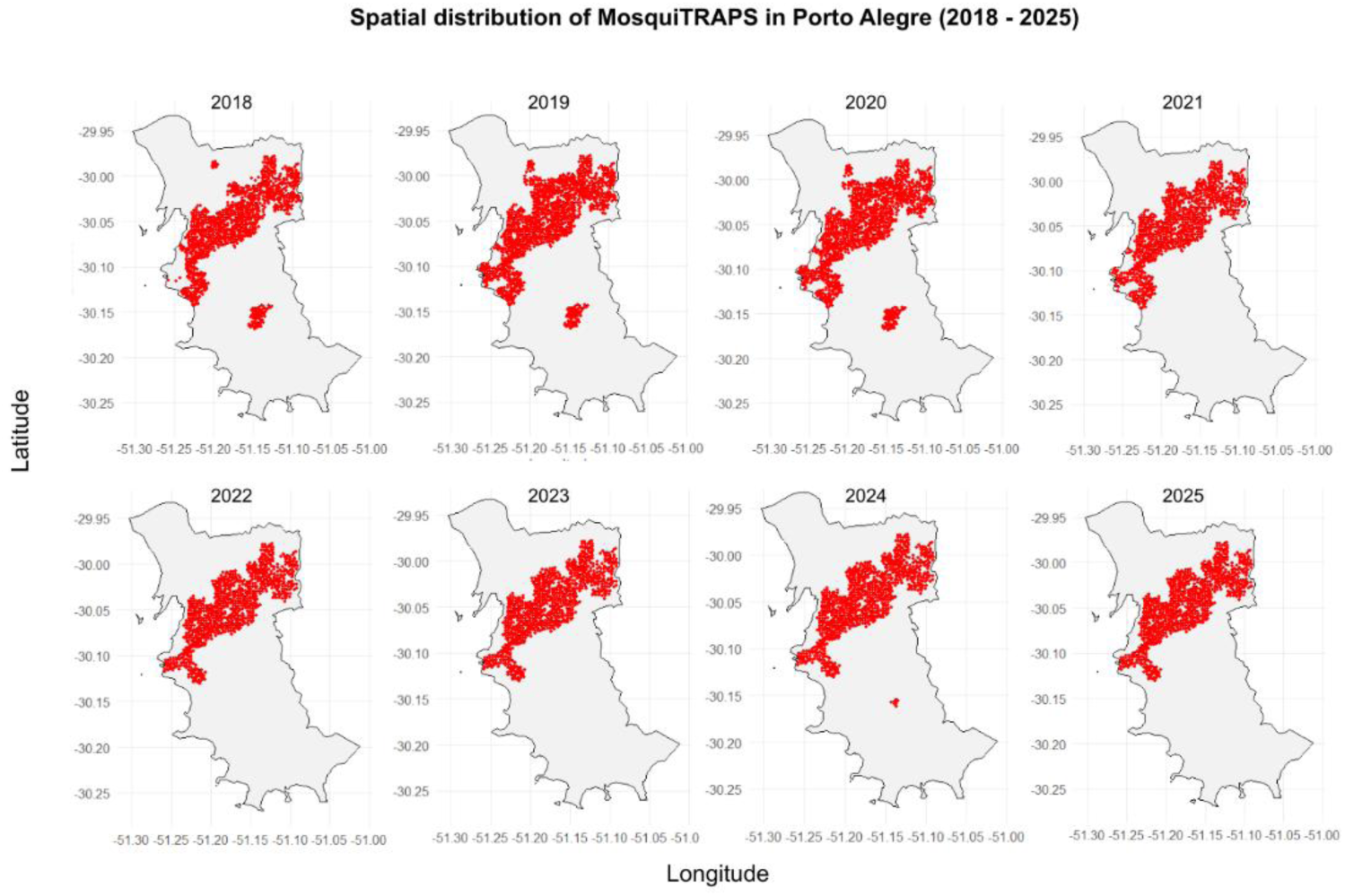
Spatial location of MosquiTRAPs in Porto Alegre from 2018 to 2025. Data were obtained from the latitude and longitude values of each distinct trap.

**S2 Fig.**
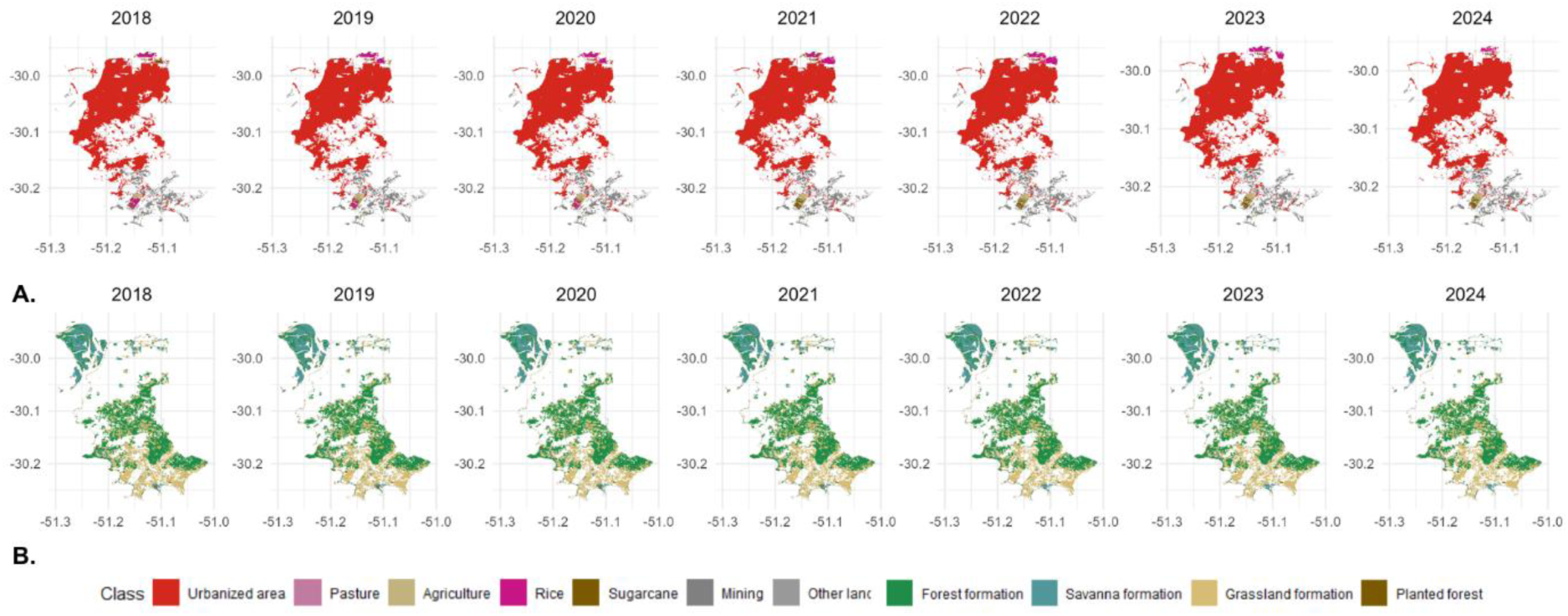
Land cover of Porto Alegre, Southern Brazil. All the data and classes were obtained from Mapbiomas Brasil. Urbanized area, Pasture, Agriculture, Rice, Sugarcane, Mining and Other classes are anthropic land use. Forest formation, Savana formation, Grassland formation and Planted forest classes are vegetation land use.

**S3 Fig.**
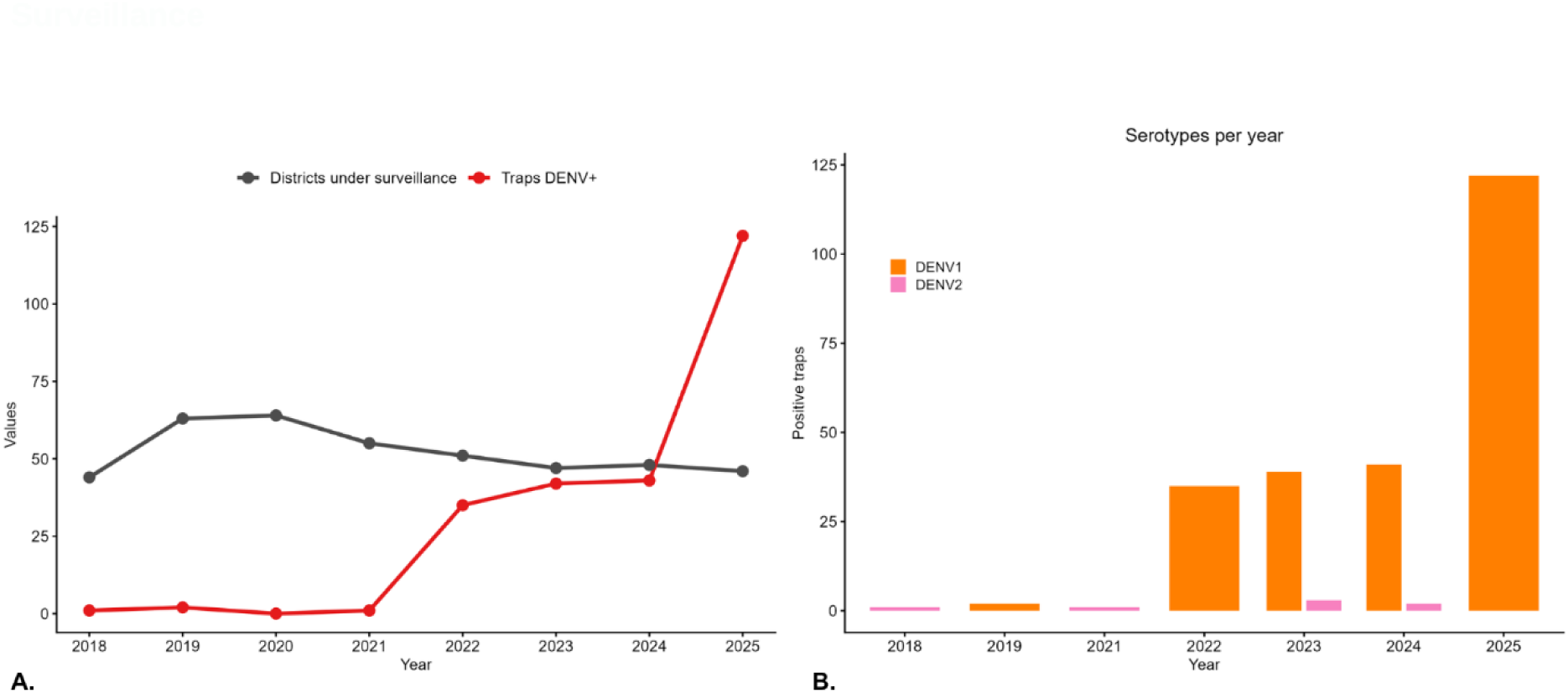
MosquiTrap dengue virus surveillance in Porto Alegre. (a) Gray line stands for districts under surveillance and red line for dengue positive traps per year from 2018 to 2025. (b) dengue serotypes obtained from positive traps per year from 2018 to 2025; orange bars are referent to DENV-1 and pink ones to DENV2.

**S4 Fig.**
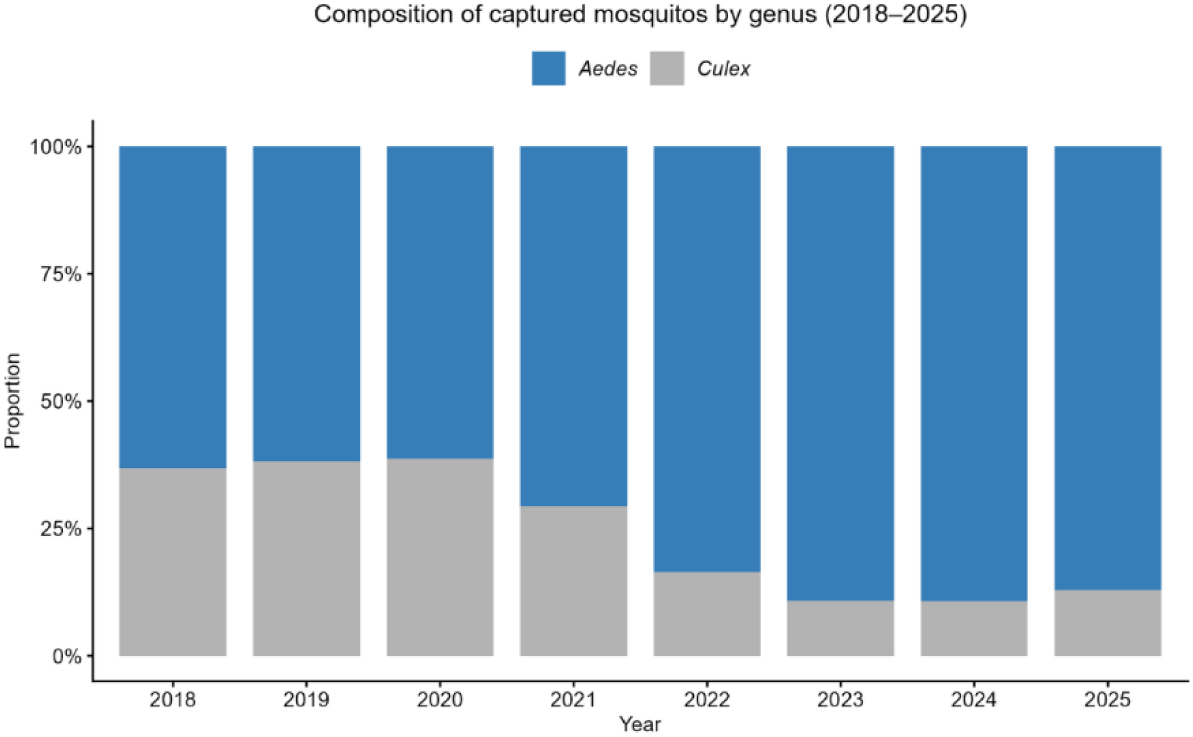
Mosquito genus captured per year from 2018 to 2025. Proportion of captured mosquitos was made by the ratio of total captured female of both mosquito genus - *Aedes* and *Culex* - among all captures.

**S5 Fig.**
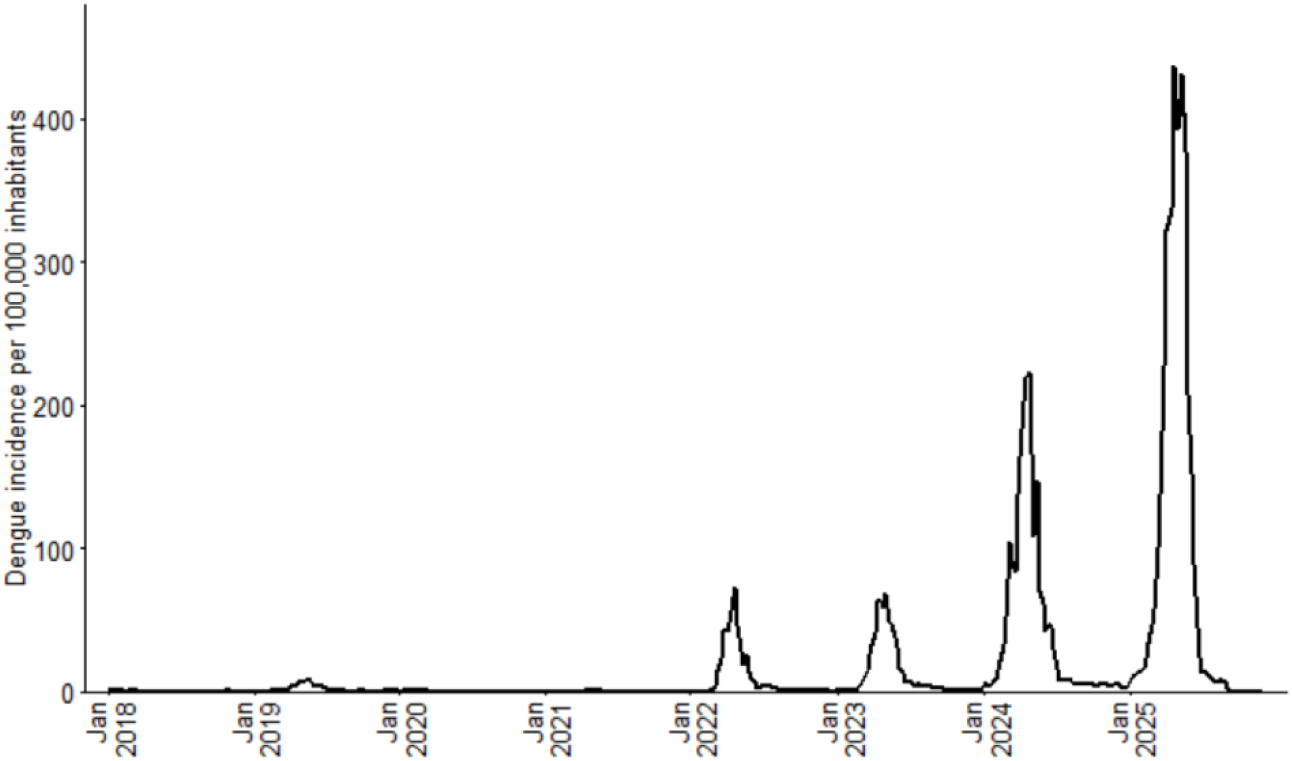
Weekly dengue incidence per 100,000 inhabitants in Porto Alegre from 2018 to 2025. Data was obtained from InforDengue Brasil.

**S6 Fig.**
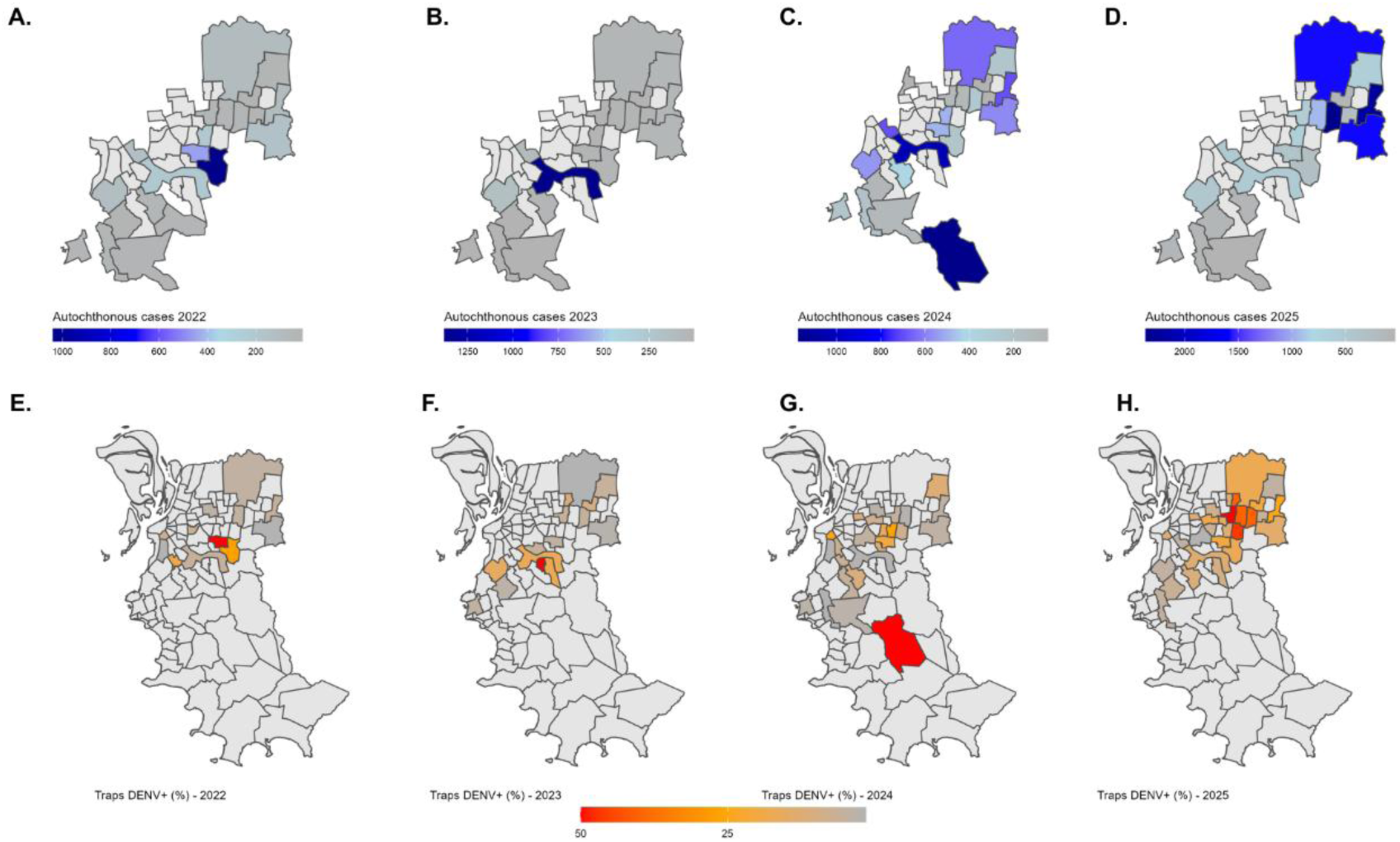
Spatial characterization of autochthonous dengue cases and DENV positive traps. Autochthonous dengue cases in Porto Alegre in 2022 (a), 2023 (b), 2024 (c) and 2025 (d). Autochthonous cases were assessed in the same districts in which positive trap analyses were performed. Proportion of dengue positive traps among total traps in 2022 (e), 2023 (f), 2024 (g) and 2025 (h).

**S7 Fig.**
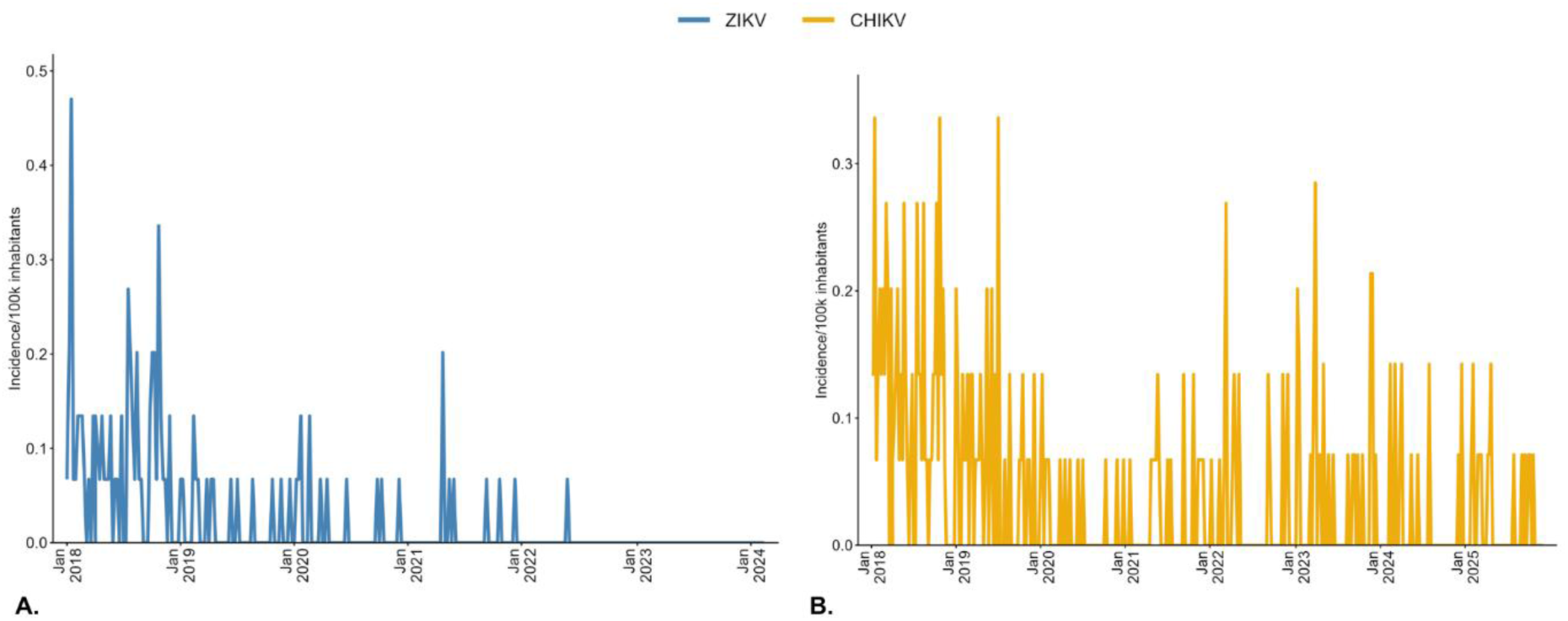
Weekly incidence of Zika virus (ZIKV) and chikungunya virus (CHIKV) in Porto Alegre, Brazil, from 2018 to 2025. Weekly incidence values of zika virus (ZIKV), yellow line (a), and chikungunya virus (CHIKV), blue line, in Porto Alegre from 2018 to 2025. Data were obtained from InfoDengue Brasil.

---

**S1 Table.** Moran’s I of mosquito infestation per year. *** p < 0.001.

| Year | *Aedes aegypti* |  | *Aedes albopictus* |  |
| --- | --- | --- | --- | --- |
|  | Moran's I | Z-score | Moran's I | Z-score |
| 2018 | 0.3717*** | 6.6302 | 0.3717*** | 4.9985 |
| 2019 | 0.5406*** | 9.4742 | 0.2859*** | 5.4709 |
| 2020 | 0.5518*** | 9.6615 | 0.3731*** | 7.1281 |
| 2021 | 0.4699*** | 8.2891 | 0.4220*** | 8.2377 |
| 2022 | 0.4900*** | 8.6716 | 0.2756*** | 5.3482 |
| 2023 | 0.6235*** | 10.91 | 0.4377*** | 9.5235 |
| 2024 | 0.5585*** | 9.796 | 0.3990*** | 8.4597 |
| 2025 | 0.5930*** | 10.399 | 0.3210*** | 6.0158 |

**S2 Table.** Kendall correlation of climate variables and mosquito infestation per district (2018 - 2025). * p < 0.05, ** p < 0.01.

| District | Variables | $\tau$ |
| --- | --- | --- |
| Boa Vista | Accumulated precipitation - *Aedes aegypti* | 0.6428* |
| Jardim Itu | Accumulated precipitation - *Aedes aegypti* | 0.7142* |
| Jardim Leopoldina | Accumulated precipitation - *Aedes aegypti* | 0.7857* |
| Jardim Sabará | Accumulated precipitation - *Aedes aegypti* | 0.6428* |
| Vila Jardim | Accumulated precipitation - *Aedes aegypti* | 0.7142** |
| Três Figueiras | Accumulated precipitation - *Aedes aegypti* | 0.7142** |
|  | Relative humidity - *Aedes aegypti* | 0.5714** |
| Mário Quintana | Accumulated precipitation - *Aedes aegypti* | 0.7857* |
| Partenon | Relative humidity - *Aedes albopictus* | 0.6428* |
| Sarandí | Accumulated precipitation - *Aedes albopictus* | 0.6428* |
| Teresópolis | Mean temperature - *Aedes albopictus* | 0.7142* |
|  | Relative humidity - *Aedes albopictus* | 0.6428* |
| Glória | Mean temperature - *Aedes albopictus* | 0.5714* |
|  | Relative humidity - *Aedes albopictus* | 0.6428* |

**S3 Table.** Moran’s I of dengue cases per year. ** p < 0.01.

| Year | Autochthonous cases |  |
| --- | --- | --- |
|  | Moran's I | Z-score |
| 2025 | 0.1861** | 3.4954 |

**S4 Table.** Moran’s I of DENV+ traps per year. ** p < 0.01, *** p < 0.001.

| Year | DENV+ traps |  |
| --- | --- | --- |
|  | Moran's I | Z-score |
| 2022 | 0.1384*** | 3.2264 |
| 2023 | 0.3085*** | 7.4082 |
| 2024 | 0.1445** | 2.9429 |
| 2025 | 0.5521*** | 10.044 |

**S5 Table.**
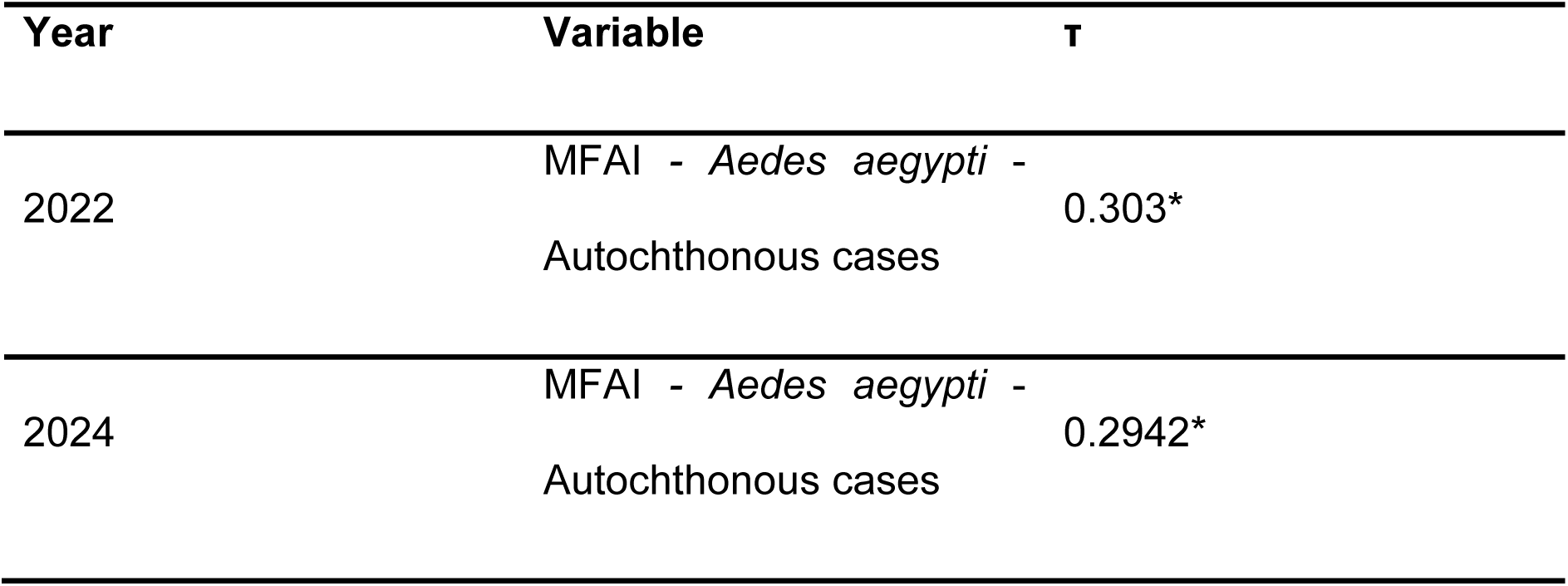
Correlation of *Aedes* proportion and autochthonous cases per year. ** p < 0.01, *** p < 0.001.

**S6 Table.** LASSO coefficients for MFAI *Aedes* model and dengue incidence model.

| LASSO |  |  |
| --- | --- | --- |
| MFAI - *Aedes aegypti* | MFAI - *Aedes albopictus* | Dengue incidence per 100,000 inhabitants |

| Variable | Coefficient | Variable | Coefficient | Variable | Coefficient |
| --- | --- | --- | --- | --- | --- |
| (Intercept) | -3,08491 | (Intercept) | -0,1436 | (Intercept) | 0,6559 |
| Precipitation (mm) | -0,00092 | Precipitation (mm) | -0,00008 | MFAI – *Aedes aegypti* | 0,66421 |
| Temperature (°C) | -0,00131 | Temperature (°C) | 0 | VFII - *Aedes aegypti* | -0,04597 |
| Humidity (%) | 0,00758 | Humidity (%) | 0,00034 | MFAI - *Aedes aegypti* (lag 1) | 0,7273 |
| Precipitation (lag 1) | -0,003395 | Precipitation (lag 1) | -0,00012 | MFAI - *Aedes aegypti* (lag 2) | 0,8393 |
| Precipitation (lag 2) | -0,00339 | Precipitation (lag 2) | -0,00013 | MFAI - *Aedes aegypti* (lag 3) | 0,6883 |
| Precipitation (lag 3) | -0,00217 | Precipitation (lag 3) | -0,000039 | MFAI - *Aedes aegypti* (lag 4) | 1,2978 |
| Precipitation (lag 4) | -0,00067 | Precipitation (lag 4) | -0,000042 | VFII - *Aedes aegypti* (lag 1) | -0,2053 |
| Temperature (lag 1) | 0,01955 | Temperature (lag 1) | 0,0004 | VFII - *Aedes aegypti* (lag 2) | -0,0458 |
| Temperature (lag 2) | 0,00932 | Temperature (lag 2) | 0,0006 | VFII - *Aedes aegypti* (lag 4) | -0,0209 |
| Temperature (lag 3) | 0,22556 | Temperature (lag 3) | 0,0008 | MFAI – *Aedes albopictus* | -10,3946 |
| Temperature (lag 4) | 0,04078 | Temperature (lag 4) | 0,001 | VFII - *Aedes albopictus* | 2,1105 |
| Humidity (lag 1) | 0,00858 | Humidity (lag 1) | 0,00039 | MFAI - *Aedes albopictus* (lag 1) | -9,055 |
| Humidity (lag 2) | 0,00721 | Humidity (lag 2) | 0,00033 | MFAI - *Aedes albopictus* (lag 2) | -8,928 |
| Humidity (lag 3) | 0,0086 | Humidity (lag 3) | 0,00021 | MFAI - *Aedes albopictus* (lag 3) | -9,8462 |
| Humidity (lag 4) | -0,0041 | Humidity (lag 4) | 0,0007 | MFAI - *Aedes albopictus* (lag 4) | -8,7928 |
|  |  |  |  | VFII - *Aedes albopictus* (lag 1) | 1,281 |
|  |  |  |  | VFII - *Aedes albopictus* (lag 2) | 1,294 |
|  |  |  |  | VFII - *Aedes albopictus* (lag 3) | 0,443 |
|  |  |  |  | VFII - *Aedes albopictus* (lag 4) | 1,2292 |
|  |  |  |  | IndexP | -0,363 |
|  |  |  |  | IndexP (lag 1) | -0,0211 |
|  |  |  |  | IndexP (lag 2) | -0,021 |
|  |  |  |  | IndexP (lag 3) | -0,037 |
|  |  |  |  | IndexP (lag 4) | -0,0479 |

## Notes

### Competing Interest Statement

The authors have declared no competing interest.

### Funding Statement

This study did not receive any funding.

### Author Declarations

Ethics Committee of the Municipal Health Department of Porto Alegre (CEP SMSPA) approved this work.

